# Reliability and limits of transport-ventilators to safely ventilate severe patients in special surge situations

**DOI:** 10.1101/2020.10.07.20208561

**Authors:** Dominique Savary, Arnaud Lesimple, François Beloncle, François Morin, François Templier, Alexandre Broc, Laurent Brochard, Jean-Christophe Richard, Alain Mercat

## Abstract

**Background:** Several Intensive Care Units (ICU) have been overwhelmed by the surge of COVID-19 patients thus necessitating to extend ventilation capacity outside the ICU where air and oxygen pressure are not always available. Transport ventilators requiring only O_2_ source may be used to deliver volume-controlled ventilation.

**Objective:** To evaluate the performances of four transport ventilators compared to an ICU ventilator simulating severe respiratory conditions.

**Materials and methods:** Two pneumatic transport ventilators, (Oxylog 3000, Draeger; Osiris 3, Air Liquide Medical Systems) and two turbine transport ventilators (Elisee 350, ResMed; Monnal T60, Air Liquide Medical Systems) were compared to an ICU ventilator (Engström Carestation – GE Healthcare) using a Michigan training test lung. We tested each ventilator with different set volumes Vt_set_ (350, 450, 550 ml) and different compliances (20 or 50 ml/cmH_2_O) and a resistance of 15 cmH_2_ 0/L/sec based on values recently described in COVID-19 Acute Respiratory Distress Syndrome. Volume error was measured, as well as the trigger time delay during assist-control ventilation simulating spontaneous breathing activity with a P_0.1_ of 4 cmH_2_0.

**Results:** Grouping all conditions, the volume error was 2.9 ± 2.2 % for Engström Carestation; 3.6 ± 3.9 % for Osiris 3; 2.5 ± 2.1 % for Oxylog 3000; 5.4 ± 2.7 % for Monnal T60 and 8.8 ± 4.8 % for Elisee 350. Grouping all conditions, trigger delay was 42 ± 4 ms, 65 ± 5 ms, 151 ± 14 ms, 51 ± 6 and 64 ± 5 ms for Engström Carestation, Osiris 3, Oxylog 3000, Monnal T60 and Elisee 350, respectively.

**Conclusions:** In special surge situations such as COVID-19 pandemic, most transport ventilators may be used to safely deliver volume-controlled ventilation in locations where only oxygen pressure supply is available with acceptable volume accuracy. Performances regarding triggering function are generally acceptable but vary across ventilators.

## Introduction

During the COVID 19 pandemic, some hospitals experienced the greatest shortage of ventilators ever seen since the heroic times of the polio epidemic in the 1950s. In this context, different alternative solutions including ventilator sharing and use of anesthesia ventilators have been considered to manage intubated patients with severe lung failure outside the walls of the ICU [1,2]. For this purpose, ventilators must be relatively easy for the users, able to deliver reliable volume-controlled ventilation in difficult mechanical conditions adapted to the principles of protective lung ventilation. Importantly, they must allow to vary FiO_2_ without requiring two pressurized sources of gas, (i.e. wall air and oxygen at 50 psi). Several transport ventilators are based on pneumatic systems and Venturi systems for gas mixing; others use an internal turbine for pressurization; in this case, a low pressure source of oxygen is needed for increasing oxygen concentration, but not as a pressure source. They all represent interesting solutions in this context and could fulfill the requirements mentioned. Pneumatic transport ventilators have been used for decades both for in- and out-of-hospital transport. Their robustness and their relative technological simplicity could potentially facilitate massive industrial production. The general view on these ventilators is, however, that their limitations make them acceptable for a short period like transport but make them incompatible with the safe delivery of difficult ventilation for very sick patients over prolonged periods. Undoubtedly, they have limited capacities regarding ventilation modes and monitoring, but we questioned whether their performance for delivering lung protective ventilation in patients with ARDS merited to be tested again with these objectives in mind. Indeed, discarding their use in a context of surge could limit the extension of offering ICU beds possibilities for mechanically ventilated patients. The performance and limits of these ventilators have not been specifically tested with the appropriate settings and realistic conditions simulating the respiratory mechanics of patients with COVID-19 induced ARDS [3-6].

The aim of the present study was to evaluate the safety, reliability and limitations of ventilation provided by these different technologies in simulated bench conditions of severe respiratory mechanics mimicking patients with COVID-19 induced ARDS.

## Materials and Methods

### 1. Ventilators

#### Brands

Four portable ventilators necessitating only one O_2_ pressurized gas source were included in the study. Two pneumatic transport ventilators using Venturi systems to mix air to oxygen were tested: the Oxylog 3000 (Draeger, Lubeck, Germany) and the Osiris 3 (Air Liquide Medical Systems, Antony, France). Two turbine transport ventilators necessitating additional oxygen only to increase FiO_2_ were also tested: the Elisee 350 (ResMed, San-Diego, USA) and the Monnal T60 (Air Liquide Medical Systems, Antony France). The objective was to assess their capability to deliver acceptable ventilation by comparing their performances to a standard ICU ventilator: Engström Carestation (GE healthcare, Madison, USA). The characteristics of the five ventilators are given in Table 1.

**Table 1.** General characteristics of the ventilators.

|  | **Engström**<br>**Carestation** | **Osiris 3** | **Oxylog 3000** | **Monnal T60** | **Elisee 350** |
| --- | --- | --- | --- | --- | --- |
| *Manufacturer* | GE Healthcare | Air Liquide Medical<br>Systems | Draeger | Air Liquide Medical<br>Systems | Resmed |
| *Weight [kg]* | 31.0 | 5.0 | 5.4 | 3.7 | 4.0 |
| *Working pressure* | Pressurized oxygen<br>and air | Pressurized oxygen | Pressurized oxygen | Pressurized oxygen | Pressurized oxygen |
| *Expired volume*<br>*monitoring* | Yes | Yes | Yes | Yes | Yes |
| *Tidal volume (Vt) [mL]* | 20 - 2000 | 100 - 2000 | 50 - 2000 | 20 - 2000 | 50 - 2500 |
| *PEEP [cmH2O]* | 1 - 50 | 0 - 15 | 0 - 20 | 0 - 20 | 0 - 25 |
| *Peak inspiratory*<br>*pressure [cmH2O]* | 7 - 100 | 10 - 80 | PEEP + 3 - PEEP + 55 | 0 - 80 | 0 - 100 |
| *FiO<sub>2</sub> [%]* | 21 - 100 | 70 or 100 | 40 - 100 | 21 - 100 | 21 - 100 |
| *Battery duration [h]* | 0.5 - 2 | 6 - 14 | 4 | 2.5 - 5 | 3 - 6 |

#### Working principle and settings

In the two pneumatic transport ventilators tested (Oxylog 3000 and Osiris 3), the working pressure that generates ventilation comes from the high-pressure oxygen supply. These ventilators based on “Venturi-distributor” technology work as flow generator.

With the Oxylog 3000, the air-O_2_ mixing is regulated from 40% to 100% via a Venturi system coupled with a proportional inspiratory valve that also permits to directly set the volume (Vt_set_). The inspiratory flow depends on both the respiratory rate (RR) and the Inspiratory:Expiratory (I:E) ratio. In other words, for a given set volume, changing RR and/or I:E ratio keep the set Vt but affects inspiratory flow. The monitoring of the expired Vt is available via a specific flow sensor inserted between the endotracheal tube and the patient circuit.

With the Osiris 3, the pneumatic system is based on the Venturi effect to entrain ambient air into the ventilator for FiO_2_ management coupled with a distributor for inspiratory flow setting. Only two positions are available for FiO_2_: 100% or an FiO_2_ around 70%. For a given combination of I:E ratio and respiratory rate, the Vt is set by directly adjusting a Vt knob that regulates the inspiratory flow. The monitoring of the expired Vt is available via a specific flow sensor inserted between the endotracheal tube and the patient circuit.

The Elisee 350 and T60 are two turbine-based ventilators which need oxygen only to adjust FiO_2_. On those ventilators, the Vt and the inspiratory flow are directly set. Changing the respiratory rate does not affect neither Vt nor inspiratory flow.

The Engström Carestation is a classical high-quality ICU ventilator requiring two sources of pressurized gas for oxygen and air (usually wall pressure at 50-55 psi).

Performances during volume control (VC) ventilation and assist volume control ventilation (ACV) were evaluated with different conditions of simulated respiratory mechanics reproducing patients with COVID-19 induced ARDS. All experiments have been performed in the Ventilatory Laboratory (Vent-Lab) of the Angers Hospital, medical ICU.

### 2. Volume delivered and PEEP with different respiratory mechanics

We assessed the volume effectively delivered (Vte_measured_) by the ventilators in different conditions of respiratory mechanics simulated on a Michigan test lung (Michigan Instruments, Kentwood, MI, USA). A linear pneumotachograph (PNT 3700 series, Shawnee, USA) and a pressure transducer (SD160 series: Biopac systems, Goleta, CA, USA) were used to measure flow and airway pressure. Signals were converted with an analog digital converter (MP150; Biopac systems, Goleta, CA, USA) at a sample rate of 200Hz, and stored in a laptop using a dedicated software (Acknowledge, Biopac Systems). Volume was obtained from numerical integration of the flow signal.

Three set volumes (Vtset) were tested: 350 mL, 450 mL and 550 mL, which approximately cover 6ml/kg of Predicted Body Weight (PBW) for most adult patients. The different respiratory mechanics conditions tested were: compliance of 50 ml/cmH_2_O and 20 ml/cmH_2_O, both combined with a resistance of 15 cmH_2_O/L/s. The combinations of compliance and resistance tested were based on recently described COVID-19 respiratory mechanics [3-6].

Assist Control Ventilation (ACV) mode was selected and similar ventilator settings were applied for each ventilator (respiratory rate 30 cycles/min).

The pneumatic transport ventilators were set with an Inspiratory:Expiratory ratio of 1:3 (I:E) whereas a flow of 60L/min was adjusted on the Engström Carestation, Elisee 350 and Monnal T60.

The three set volumes were tested with FiO_2_ 100% and 70% (air-O_2_ mix for Osiris 3) as follows: FiO_2_ was selected, Vt_set_ was adjusted on the ventilator and Vte_measured_ was recorded and averaged over 5 cycles after stabilization. FiO_2_ was measured on Osiris 3 when air-O_2_ mix was selected with a PF300 gas analyzer (IMT Medical, Buchs, Switzerland) in different conditions (Vt = 350 - 450 - 550 mL and Compliance = 20 - 50 mL/cmH_2_O).

The performances of Venturi-based ventilation in terms of volume delivery could be altered by set inspiratory flow values [7]. To assess the impact of the inspiratory flow, Vte_error_ was calculated on the Osiris 3 ventilator. We set a volume of 450 ml using a wide range of inspiratory flows achieved by changing respiratory rate (RR). ACV mode with air-O_2_ mix was selected, a resistance of 15 cmH_2_O/L/s and a compliance of 20ml/cmH_2_O were applied and we set a I:E ratio of 1:3. The lowest RR (6 cycles/min) was chosen and was progressively increased by 4 cycles/min until reaching the maximum RR of 40 cycles/min. Vte_set_ had to be adjusted in consequence at each RR increment to keep its value at 450 ml. Vte_error_ was estimated at each step.

Two levels of PEEP were applied (10 cmH_2_O and 15 cmH_2_O) and the accuracy of the effective PEEP (PEEP_measured_) was assessed.

#### Volume error and PEEP end-points

The relative volume error (Vte_error_), which is the difference between the effective expired volume (Vte_measured_) and the set volume (Vte_set_) was calculated and averaged as previously described over the four different conditions [8,9]:

- Resistance = 15 cmH_2_O/L/s, Compliance = 20 or 50 ml/cmH_2_O, PEEP = 10 or 15 cmH_2_O

The relative volume error was expressed in percentage and defined as follows:

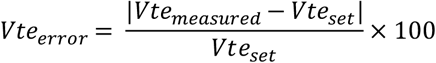

##### End-point for VT_error_

The three tidal volumes tested were chosen to cover theoretical “6 ml/kg PBW” in adult male or female patients (350, 450 and 550 ml correspond to 6ml/kg PBW for respectively 58, 75 and 92 kg PBW).

Ventilation was considered safe and acceptable when Vte_measured_ was within ± 0.5 ml/kg PBW, which covers a volume between 5.5 and 6.5 ml/kg PBW. This corresponds to an 8% difference between set and measured Vt.

##### End-point for PEEP

a difference between measured PEEP and set PEEP was acceptable when less than 2 cmH_2_O.

### 3. Synchronization and trigger performances

Assist control ventilation (ACV) with the inspiratory trigger function “on” was tested by connecting ventilators to a double chamber Michigan test lung (Michigan Instruments, Kentwood, MI, USA) to simulate spontaneous breathing. One chamber of the test lung was defined as the driving lung while the other chamber was connected to the ventilator being tested. A lung-coupling clip allowed a connection between the two chambers, so that a positive pressure created in the driving lung induced a negative pressure in the experimental lung, leading to trigger the ventilator tested.

The driving lung was connected to an Evita XL ventilator (Draeger, Lubeck, Germany) and set in volume-controlled mode with constant flow. The respiratory rate was set at 25 breaths/ min. The ventilatory settings were chosen to achieve a decrease in airway pressure 100 ms after occlusion (P_0;1_) of 4 cmH_2_0 (consistent with P_0;1_ values recently described in COVID patients [10]), measured at the airway opening of the lung model [11]. A level of PEEP was applied to the driving lung to obtain a perfect contact of the lung-coupling clip between the two chambers at the end of expiration.

For each ventilator tested, volume assist-control ventilation (ACV) mode was selected, with a tidal volume of 450 mL, a respiratory rate of 20 cycles/min and a PEEP of 10 cmH_2_O. I:E ratio was set at 1:3 for Osiris 3 and Oxylog 3000, while a flow of 60L/min was set on Elisee 350, Monnal T60 and the Engström Carestation ICU ventilator. We selected air O_2_-mix for Osiris 3, and FiO_2_ of 70% on the other ventilators. Inspiratory triggers were set at their most responsive position while avoiding auto-triggering. The trigger of the Osiris 3 was set at −0.5 cmH_2_O. Flow-triggered ventilators were set at 1 L/min for Engström Carestation and Oxylog 3000 and 2 L/min for Monnal T60 and Elisee 350.

For each configuration, trigger performance was assessed by measuring airway pressure changes using the flow trace to determine the start of inspiration [9,12]. Negative pressure drop (ΔP, cmH_2_0), Triggering Delay (TD, ms) and Pressurization Delay (PD, ms) as defined on figure 1 were computed. The overall Inspiratory Delay (ID) corresponds to the addition of TD and PD.

**Figure 1.**
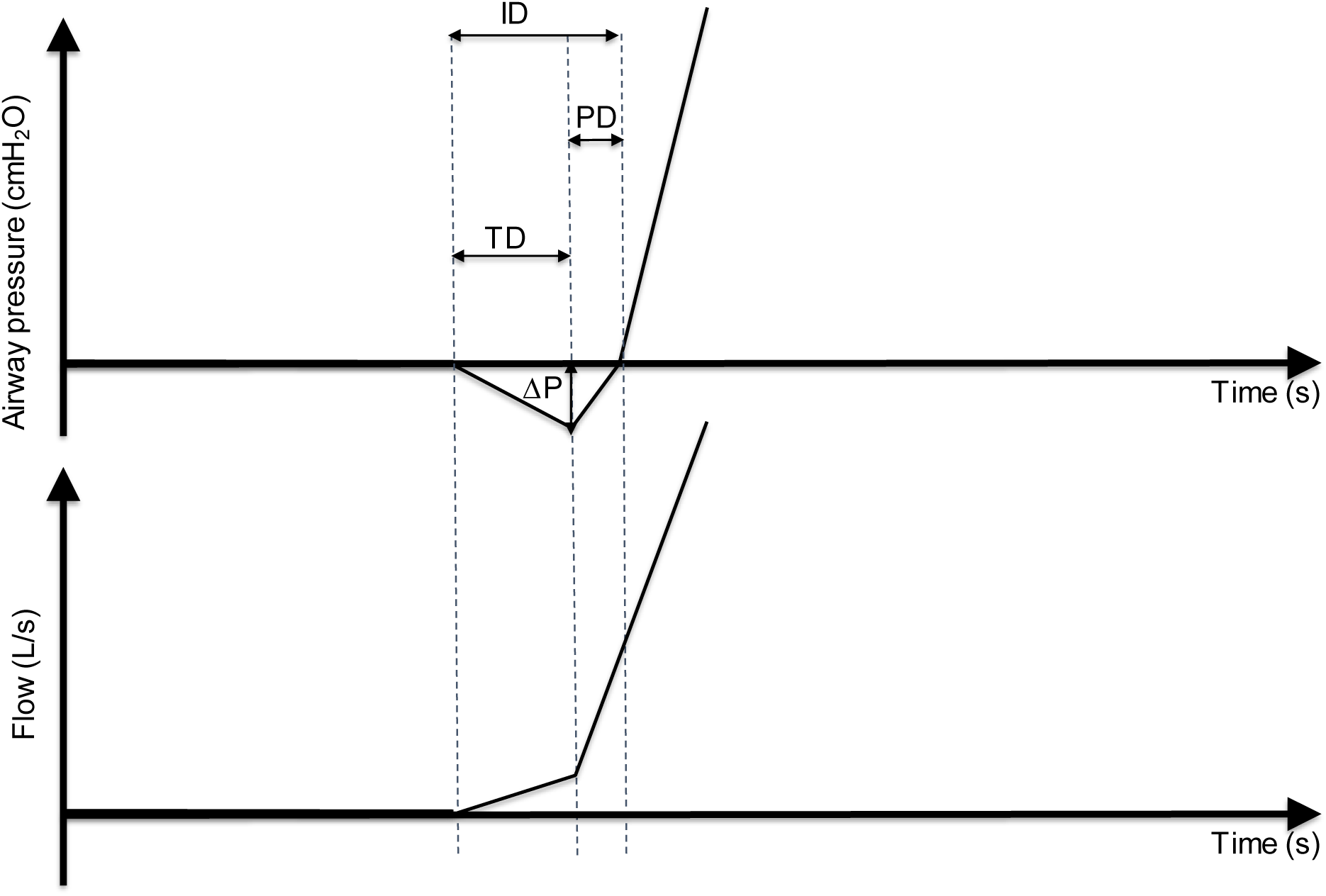
Explicative figure of ventilator triggering assessment. The figure illustrates ventilator triggering assessment. Airway pressure (Paw) and flow are displayed. Triggering delay (TD) is the delay between the onset of airway pressure drop (“patient” effort) and flow delivery by the ventilator. Pressurization delay (PD) is defined by the time at which the airway pressure comes back to the level of PEEP. The addition of TD and PD gives the inspiratory delay (ID). The drop of airway pressure (ΔP) due to patient effort is also shown on the figure.

#### End-point

triggering function was considered as “safe and acceptable” when TD was less than 100 ms.

### 4. Statistical analysis

Continuous variables were expressed as mean ± SD values averaged from 5 consecutive breaths. These variables were compared using an ANOVA test. The type I significance level was set at 0.05. When the global F was significant, post hoc tests were computed using a student t-test with Bonferroni correction, which sets the level of significance for pairwise differences between the five ventilators at 0.005.

## Results

### 1. Volume delivered and PEEP measured with different respiratory mechanics

Results are displayed in figure 2 and mean volume errors (Vte_error_) for each ventilator are shown in Table 2. When all conditions and set volumes were included, the Engström Carestation was the most precise ventilator, and the Oxylog 3000 was comparable. Each ventilator performance was considered as acceptable (delta Vt ± 0.5ml/kg PBW) except for one turbine ventilator (Elisee 350). The impact of FiO_2_ selection (FiO2 100% or 70%) on volume error was significant considering all ventilators (p < 0.05), reaching a maximum of 11.9 % for Elisee 350 with FiO_2_ 100% (changes for each ventilator are available on table 2). The impact of compliance on volume error was not significant considering all ventilators (p > 0.05) and changes for each ventilator are available on table 2. FiO_2_ measured on Osiris 3 reached 72.3 ± 1.7% regarding all the conditions tested.

**Table 2.**
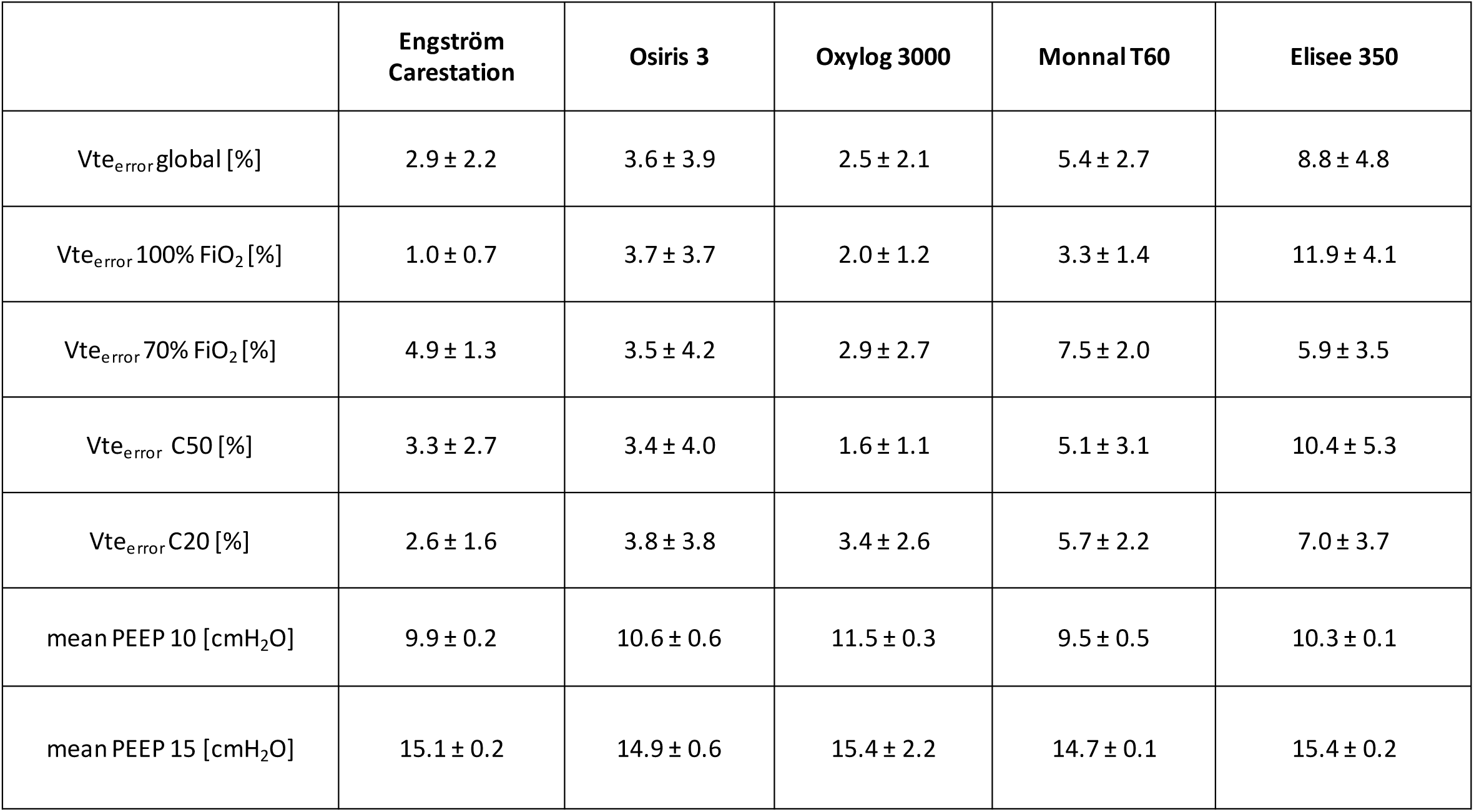
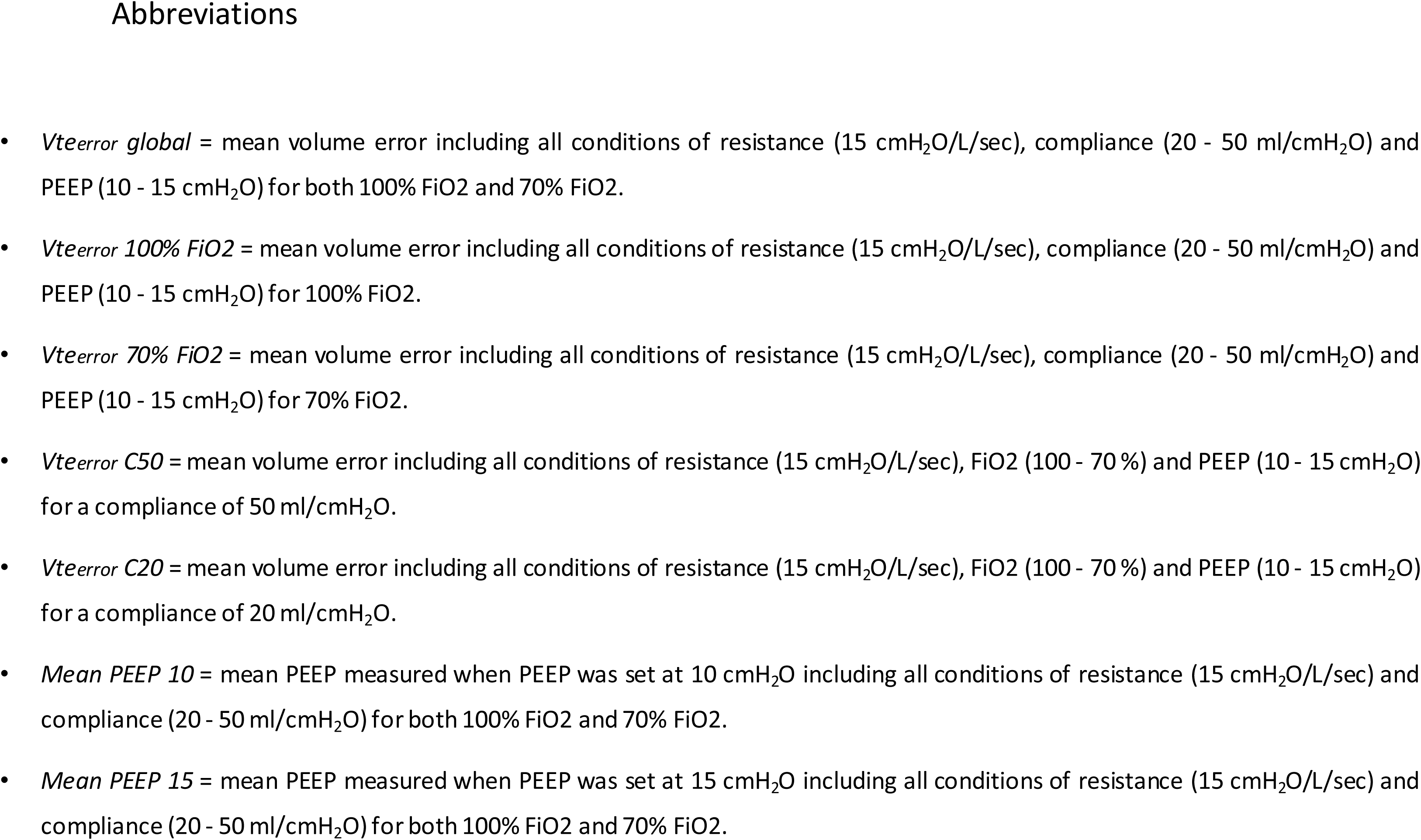
Mean volume errors and Positive End Expiratory Pressure (PEEP) measured for each ventilator.

**Figure 2.**
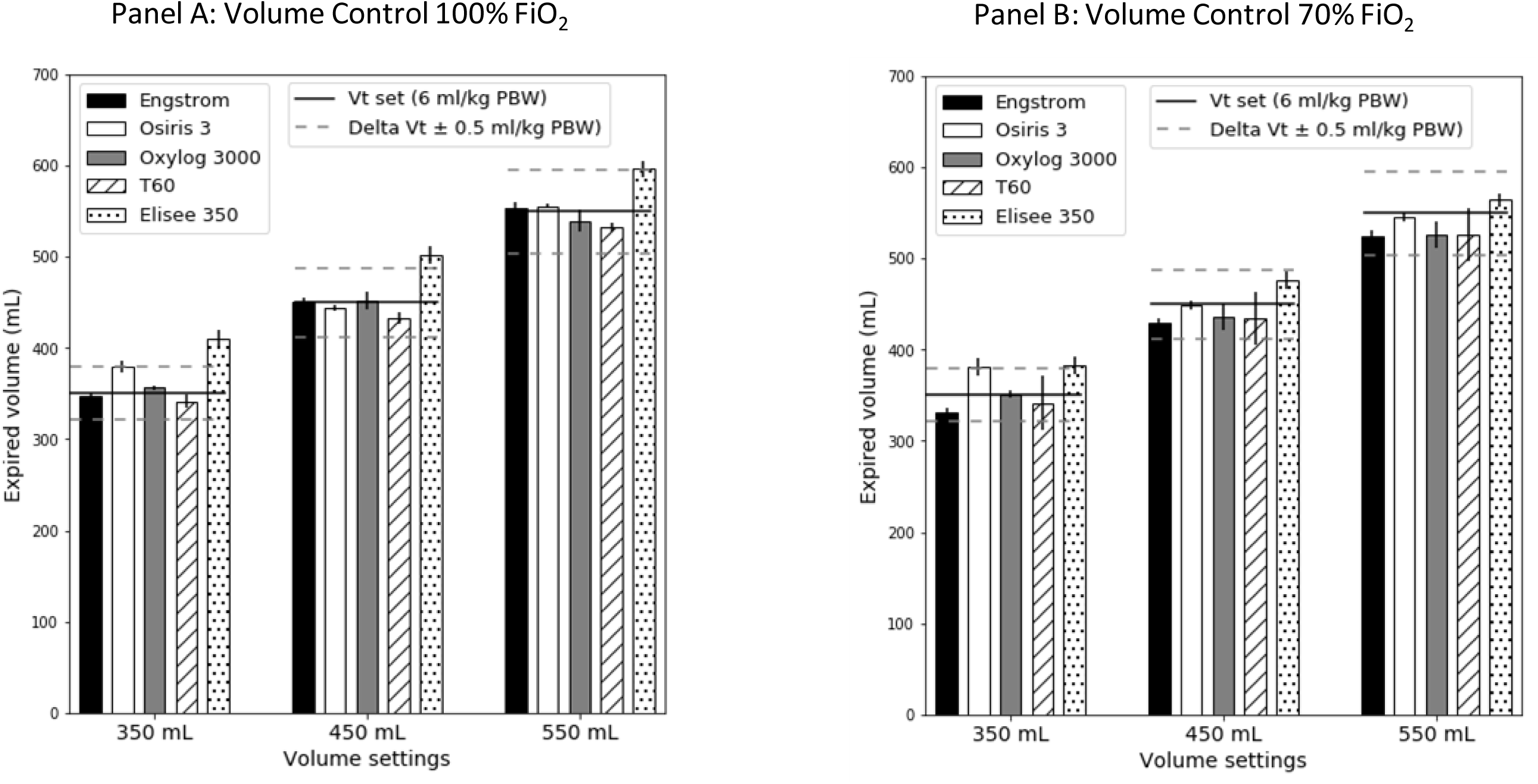
Tidal Volume delivery in volume control ventilation in static conditions. **Panel A.** The histogram represents the mean expired volumes measured for each ventilator according to the three Vt set in 100% FiO_2_. The average was computed over the four conditions of resistance (15 cmH_2_O/L/sec), compliance (20 - 50 ml/cmH_2_O) and PEEP (10 - 15 cmH_2_O). The three tidal volumes tested were chosen to cover 6 ml/kg PBW, with 350, 450 and 550 ml corresponding to 6ml/kg PBW for respectively 58, 75 and 92 kg PBW. Limits of acceptable ventilation are displayed with dotted lines and defined as a volume change within ± 0.5 ml/kg PBW, which corresponds to a Vt between 5.5 and 6.5 ml/kg PBW. **Panel B.** The histogram represents the mean expired volumes measured for each ventilator according to the three Vt set in 70% FiO_2_. The average was computed over the four conditions of resistance (15 cmH_2_O/L/sec), compliance (20 - 50 ml/cmH_2_O) and PEEP (10 - 15 cmH_2_O).

When all conditions were put together, differences between measured PEEP and set PEEP were less than 2 cmH_2_O as shown on table 2.

### 2. Impact of inspiratory flow on pneumatic ventilators

The effect of inspiratory flow rates on Vte_error_ for Osiris 3 is shown on figure 3. The lowest values of inspiratory flow are associated with a Vte_error_ above 8% (delta Vt ± 0.5ml/kg PBW). Performances are acceptable when inspiratory flow (resulting from the combination of Vt, I:E ratio and respiratory rate) was strictly above 30 L/min, which corresponds to a respiratory rate higher than 20 cycles/min.

**Figure 3.**
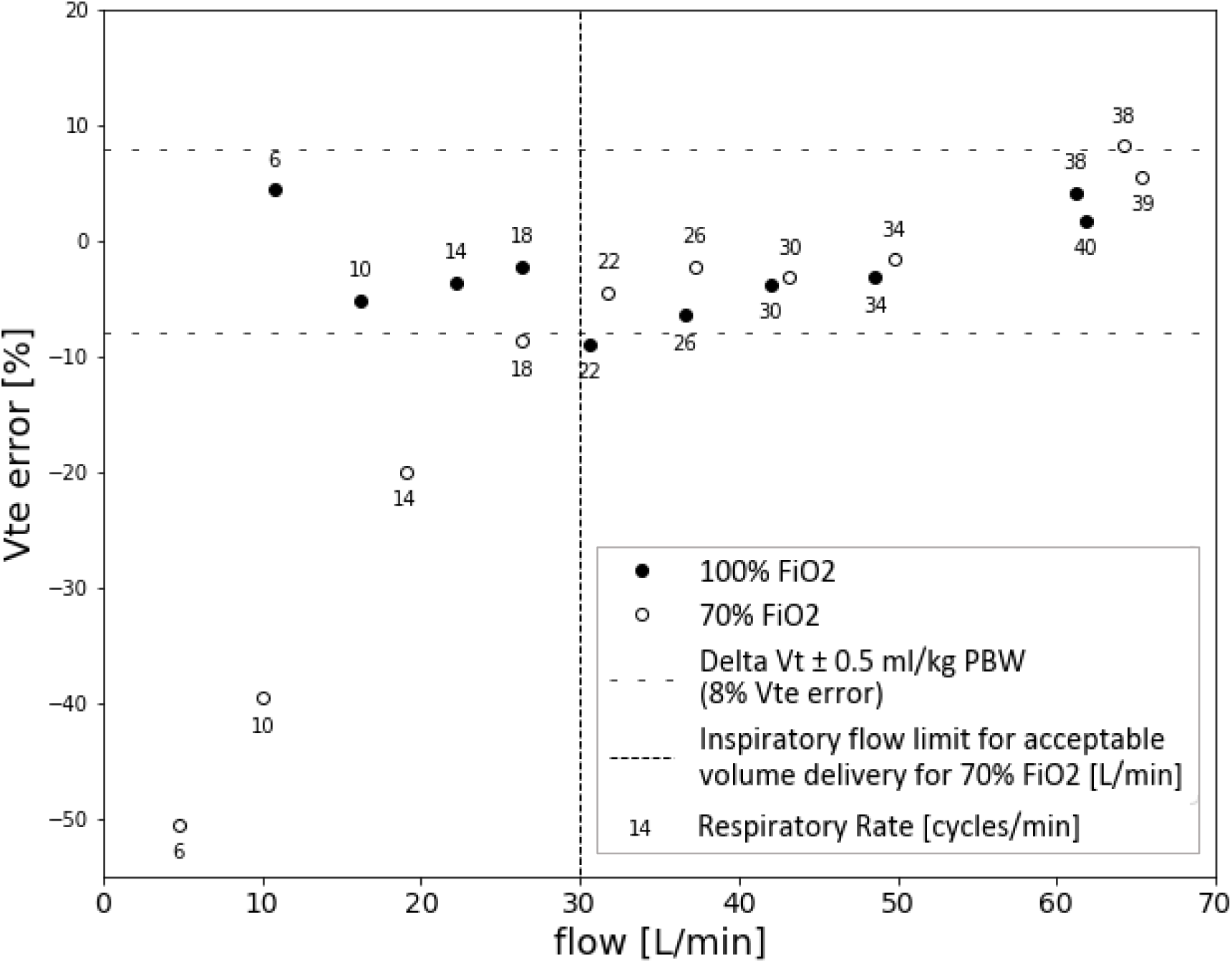
Impact of flow on effective volume with Osiris 3 ventilator. This figure shows the volume error of the Osiris 3 expressed in % of Vt set according to different inspiratory flows obtained at a constant 450 ml Vt set. Compliance, resistance and PEEP were set at 20 ml/cmH_2_O, 15 cmH_2_O/L/sec and 10 cmH_2_O respectively. Black circles were obtained with 100% FiO_2_ while the white circles were obtained with 70% FiO_2_. Respiratory rate associated with each point is also displayed. This figure illustrates that for an inspiratory flow below 30 L/min, the Vt error is substantial with 70% FiO_2_. The Vt error is within ± 0.5 ml/kg PBW (which corresponds to an 8% difference between set and measured Vt) whatever the inspiratory flow when 100% FiO_2_ is selected.

### 3. Synchronization and trigger performances during ACV

Inspiratory trigger was evaluated for each ventilator and results are displayed on figure 4. All simulated efforts triggered a ventilatory cycle. The Triggering Delay was 42 ± 4 ms, 65 ± 5 ms, 151 ± 14 ms, 51 ± 6 ms and 64 ± 5 ms for Engström Carestation, Osiris 3, Oxylog 3000, Monnal T60 and Elisee 350, respectively (all conditions grouped). Each ventilator performance was considered as acceptable (TD < 100ms) except for one pneumatic ventilator (Oxylog 3000).

**Figure 4.**
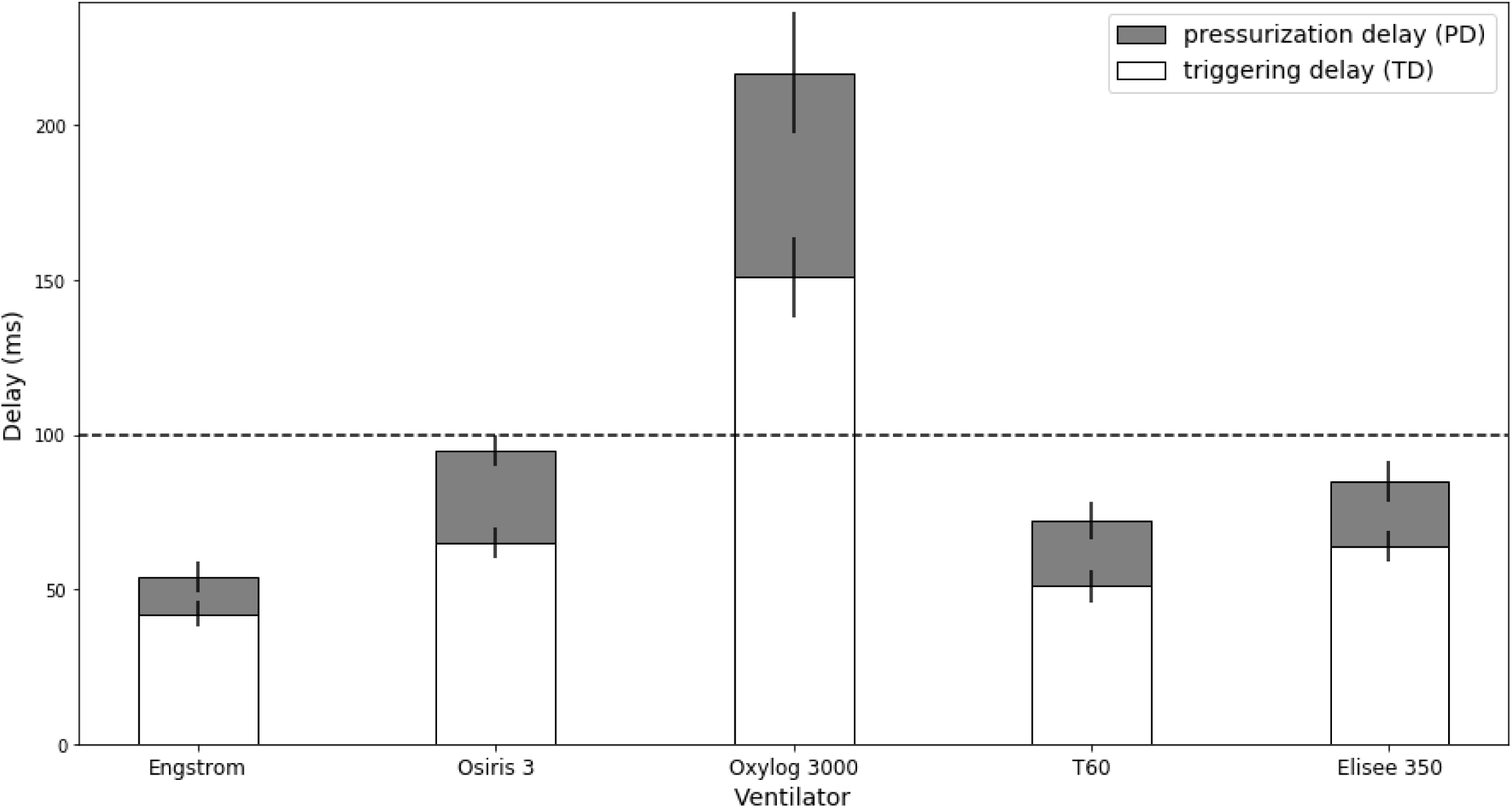
Triggering efficiency in volume assist-control ventilation. The figure illustrates the triggering efficiency for each ventilator tested during assist-control ventilation using the Michigan test lung to simulate spontaneous breathing. A PEEP of 10 cmH_2_O, a compliance of 20 and 50 ml/cmH_2_O and a resistance of 15 cmH_2_O/L/sec were selected. Triggering Delay (TD, ms) and Pressurization Delay (PD, ms) were computed. A definition of TD and PD is available on figure 1. Triggering function was considered safe and acceptable when TD was less than 100 ms.

The Inspiratory Delay (ID) was measured at 54 ± 5 ms for Engström Carestation, 95 ± 5 ms for Osiris 3, 217 ± 21 ms for Oxylog 3000, 72 ± 6 ms for Monnal T60 and 85 ± 7 ms for Elisee 350 and (p < 0.05; pairwise differences between ventilators were all significant with a p-value < 0.005). A similar trend was observed for triggering delay.

The airway pressure drop was much larger for Oxylog 3000 (−4.1 ± 0.3 cmH_2_O), than for the others:-0.9 ± 0.3 cmH_2_O for Engström Carestation, −1.9 ± 0.1 cmH_2_0 for Osiris 3,, −0.6 ± 0.1 cmH_2_O for Monnal T60 and - 0.9 ± 0.1 for Elisee 350 (p < 0.05; pairwise differences between ventilators were all significant with a p-value < 0.005, except between Osiris 3 and Elisee 350).

## Discussion

The results of the present bench test study comparing turbine and pneumatic transport ventilators to an ICU ventilator, can be summarized as follows: 1. Turbine ventilators show performance in VC and ACV very close to the ICU ventilator tested for most of the settings. 2. In severe respiratory mechanics conditions, the volume error exceeds 0.5 ml/kg PBW only for one turbine ventilator. 3. Inspiratory trigger reactivity is less than 100 ms except for one pneumatic transport ventilator. 4. Volume error delivered by the simplest pneumatic ventilator significantly increases when inspiratory flow is less than 30 L/min indicating a technological limit of the Venturi system.

The increasing number of patients requiring mechanical ventilation in the context of the COVID-19 worldwide crisis, and the ventilators shortage reported in some severely affected countries, has led to discuss the possibilities to manage intubated patients outside the walls of the ICU [2]. According to this dire scenario, simple and easy to set ventilators that only require one oxygen pressure source to function and able to deliver lung protective ventilation could be considered. In addition, an assisted volume mode that ensures the set Vt with PEEP up to 15 cmH_2_O and FiO_2_ up to 100% is required to manage high elastic load and severe shunt that characterize potentially severe COVID-19 ARDS [1,2]. Recent turbine transport and emergency ventilators have performances which are very close to conventional ICU ventilators [13,14]. In the context of “mass casualty”, as experienced with the COVID-19 crisis, pneumatic transport ventilators could be used to extend the possibility to manage intubated patients in case of ICU beds shortage. The working principle of these pneumatic ventilators is based on a “Venturi system” which is a simple technological solution that permits to manage ventilation generated by the oxygen pressurized source when a position called air-O_2_ mix is selected. Interestingly, the simplicity of such pneumatic systems permits to consider massive industrialization faster and at a lower cost. On the opposite, the Venturi system explains the limits observed with low inspiratory flow previously described with this technology [7].

### Performances during controlled ventilation

In case of high impedance, a low inspiratory flow may increase significantly volume error when the O_2_ air mix position is selected “on”. In turn, manipulating I:E ratio, respiratory rate and increasing inspiratory flow above 30 L/min permits to reverse the Vt error that is directly explained by the working principle of this ventilator (see figure 3). The technological adaptations available on Oxylog 3000 (Venturi coupled with proportional inspiratory valve) solve this problem while expired Vt monitoring available on Osiris 3 simplifies settings adaptation if required. Previous bench test studies have reported a Vt error with pneumatic basic transport ventilators that reached 20% of set Vt with resistive load [7,15]. These experiments were performed with very low set inspiratory flow thus explaining the Vt reduction observed. For this purpose, an essential recommendation is to set the I:E ratio at 1:3 (minimal available value), and set the RR above 20 /min before adjusting the knob (that controls the inspiratory flow) to reach the set Vt as indicated on the ventilator. With these recommendations, volume error measured on pneumatic transport ventilators at low compliance are closed from turbine performances and acceptable since it did not exceed 0.5 ml/kg PBW.

Of note, only the ICU and turbine ventilators tested compensate for the loss in Vt due to the compression of gas inside the circuit. Nevertheless, this effect previously quantified in ICU ventilators with inspiratory-expiratory circuits is significantly less in basic transport ventilators since they are equipped with a single limb circuit [8]. Of note, an HEPA filter can be easily adjusted on the expiratory limb and they should not expose to higher risks of viral contamination.

### Performances during assisted ventilation

Recent experience with COVID-19 induced ARDS reports that these patients often exhibit high respiratory drive and asynchrony that may require deep sedation and sometimes paralysis [4]. We therefore evaluated the behavior of these two pneumatic transport ventilators during triggered breaths since performances of their trigger have been questioned [14,15]. The synchronization was overall respected except for the Oxylog 3000 exhibiting the poorest triggering performances. The triggering time delay was consistently longer in pneumatic ventilators but acceptable except on the Oxylog 3000, compared to the ICU ventilator [14].

## Limitations

The results obtained in vitro necessitate some caution to be translated to the clinical practice, but previous studies showed that this type of simulation predicts the results observed in clinical situations with a high fidelity [8,16,17]. The lung model gives the unique opportunity to compare ventilator performances according to several simulated but standardized clinical conditions. Bench experiment also permits to accurately depict and understand advantages and limits of the different ventilator’s technologies as previously done [7]. Our experiment reported performances of only two pneumatic and two turbine ventilators while several other ventilators with similar technology are available worldwide. We did not evaluate pressure support ventilation while this approach can be useful to manage weaning of COVID-19 patients. Previous studies already showed that turbine-based ventilator significantly outperform pneumatic transport ventilators during pressure mode ventilation [13,14].

## Conclusion

The present bench study suggests that turbine technologies may replace ICU ventilators to extend ICU beds where only oxygen pressure supply is available, in special surge situations such as COVID-19 crisis. Pneumatic transport ventilators provide acceptable volume accuracy even in severe simulated conditions. For this purpose, attention is required to maintain sufficient inspiratory flow in the Osiris 3 (or any other pneumatic ventilator using a venturi system). A monitoring of expired Vt available on the two pneumatic transport ventilators tested greatly facilitates settings. Performances regarding triggering function are non-acceptable in one of the pneumatic transport ventilator thus rendering hazardous its use in assist control ventilation.

## Data Availability

All data referred in the manuscript can be provided by the authors on demand.

## Notes

### Competing Interest Statement

DS reports grants from Fisher and Paykel and travel fees from Air Liquide Medical Systems. JCR reports part time salary for research activities (Med2Lab) from Air Liquide Medical Systems and Vygon. AL is PhD student in the Med2Lab and in Vent Lab in the Angers ICU and is partially funded by Air Liquide Medical Systems. AB is master student from the Telecom-Physic-Strasbourg Strasbourg University France. FB reports personal fees from Löwenstein Medical, travel fees from Draeger and research support from Covidien, GE Healthcare and Getinge Group, outside this work. AM reports personal fees from Draeger, Faron Pharmaceuticals, Air Liquide Medical Systems, Pfizer, Resmed and Draeger and grants and personal fees from Fisher and Paykel and Covidien, outside this work. LB has received research grants and/or equipment for his research laboratory from Medtronic Covidien (PAV), Air Liquide (helium, CPR), Fisher Paykel (high flow), General Electric (lung volume; ultrasound), Sentec (PtcCO2) and Philips (sleep). All other authors declare no competing interests.

### Funding Statement

This study did not receive any grant or financial support.

### Author Declarations

Non Applicable -> bench study

